# Nurses’ Satisfaction with and Demand for the National Cancer Patient Home-Care Pilot Program

**DOI:** 10.1101/2023.11.21.23298855

**Authors:** Jung Ha Kim, Jang Won Lee, Jung Yoon Kim, Yong Eun Hong, Sol Bi Jang, Woo Jeong Lee, Kyung Hee Oh, Young Ae Kim

## Abstract

In December 2021, the “National Cancer Patient Home-Care Pilot Program” was launched in South Korea. However, there were medical institutions with low participation and where services were not provided. Therefore, this study aimed to investigate the progress of, satisfaction with, and needs of nurses’ regarding a pilot program for home care of cancer patients with a stoma. Data were collected through a survey administered to nurses from November 13, 2022, to January 19, 2023; a total of 207 nurses responded. After excluding non-answers, 196 nurses were included in the final study population. The questionnaire sought information on the characteristics of the respondent, status of management of stoma patients, status of the home-care pilot program, satisfaction with the program, and needs of the nurses. The data were analyzed using descriptive statistics. A total of 196 nurses participated in the survey, of whom 42 nurses (21.4%) participated in the home-care pilot program. Thirty-five nurses (85.4 %) were satisfied with the home-care pilot program. Thirty nurses (71.4%) reported that implementation of home care had to be expanded and standards of care improved. To vitalize and expand the pilot program for home care for cancer patients with a stoma, better medical services must be provided by improving the medical fee standard. This will enable more patients and medical institutions to participate in the pilot program.

## Introduction

An ostomy is a surgically created fistula that helps individuals with impaired bowel elimination to redirect waste into an external bag [1]. In South Korea, the number of people with a permanent ostomy is recorded at 16,779 as of 2022, but this number is estimated to be higher when temporary ostomies and unregistered individuals with disabilities are taken into account [2]. The 2020 National Survey on Persons with Disabilities found that 83.6% of registered individuals with ostomy-related disabilities underwent either colostomy or ileostomy [3]. Out of all patients who underwent ostomy surgery, 80.1% underwent the procedure for cancer treatment, with 80.1% of patients with colon or rectal cancer requiring an ostomy after surgery for anus resection or in the case of bowel obstruction [4]. According to the National Cancer Registry, while the incidence of colon cancer appears to be decreasing because of early cancer screenings and advancements in medical technology, it remains the third most common cancer in South Korea. Further, the percentage of patients aged 70 and above is increasing among individuals with colon cancer, and the prevalence of colon cancer is also on the rise [5].

Patients with cancer with an ostomy may be vulnerable to physical, psychological, and social issues because of the presence of a stoma. Because of the ostomy, they cannot regulate their bowel and urinary elimination, and concerns about gas and odor may cause physical, psychological (e.g., depression), and social problems (e.g., avoidance of work and other social activities) [6]. In addition, poor stoma care, peristomal skin care, and stoma bag management may increase the risk for infection and complications [7]. The incidence of stoma complications ranges from 2.9% to 81.8%, with the most common complications being peristomal skin problems and parastomal hernias [8]. Further, early complications that may occur within 30 days of ostomy creation include stoma ischemia, necrosis, mucosal and skin detachment, and peristomal abscess, whereas late complications that may occur after a patient’s physiological adaptation include parastomal hernia, prolapse, and aneurysm [9]. Hence, inappropriate stoma care causes complications, which in turn impair patients’ quality of life and ultimately require additional medical interventions.

Therefore, continued self-care after discharge is essential. The UK Association of Stoma Care Nurses recommends educating and training patients about proper stoma care at home and emphasizes the importance of continuous follow-up care and support even after discharge [10,11]. To ensure that patients are prepared for self-care of their stoma at home, stoma nurses must educate inpatients on stoma care, including attaching and removing stoma bags and handling potential problems during stoma care, such as bleeding, proper use of stoma care products, assessment of stoma contents, and assessment of stoma and peristomal skin before discharge [1]. Further, stoma nurses must provide patients with sufficient psychological support and information about how to purchase essential products and how to handle problems related to stoma and peristomal skin [10]. However, one-time stoma care education provided to patients and caregivers during a short-term stay in the hospital does not adequately prepare patients for self-care of stomas at home [12]. According to the Korea Disabled People’s Development Institute, 80.4% of patients who underwent colostomy, ileostomy, or urostomy surgery had ostomy care education at the hospital immediately after the surgery, with 78.0% stating that they had received self-care education 1–3 times and 55.6% claiming that the stoma care education offered by the hospital was inadequate [4]. Therefore, programs that monitor and provide appropriate management and self-care instructions for patients with stomas must be developed, using diverse approaches and media [13].

A pilot project, the “National Cancer Patient Home-Care Pilot Program,” was launched in South Korea in December 2021 for the continuous monitoring of cancer patients discharged with an ostomy, including stoma care and complication prevention. However, in an interim evaluation of the program, more than 50% of the participating healthcare facilities did not submit an inspection document, and some healthcare facilities did not provide home-care services for cancer patients with an ostomy. These findings suggest that further research is needed to address these issues [14]. This study aimed to investigate the current status of the pilot program, identify participants’ satisfaction and needs, and propose measures to improve the program based on the findings to transition it to a fully operational program.

## Methods

### Overview of cancer patient home-care pilot program

Healthcare facilities equivalent to a hospital-level facility or higher are eligible to participate in National Cancer Patient Home-Care Pilot Program after forming a home-care team consisting of a surgeon, stoma nurse, and nutritionist. Education and consultation fees were divided into two: education and consultation fee I, which encompasses professional education and consultation provided by a physician to an outpatient or their caregiver regarding the characteristics of an ostomy and treatment process to help with self-care at home; and education and consultation fee II, which encompasses education and consultation provided by a home-care team to an outpatient or inpatient or their caregiver regarding the disease and health management to help them practice self-care at home, such as preventing stoma complications. The patient management fee pertains to periodic monitoring of the patient’s clinical information following discharge by the home-care team and the provision of bidirectional virtual services required for home care to patients with an ostomy. The home-care team may claim patient management fees only if they provide virtual patient management services at least twice a month using a platform that allows bidirectional communication (Fig 1).

**Fig 1.**
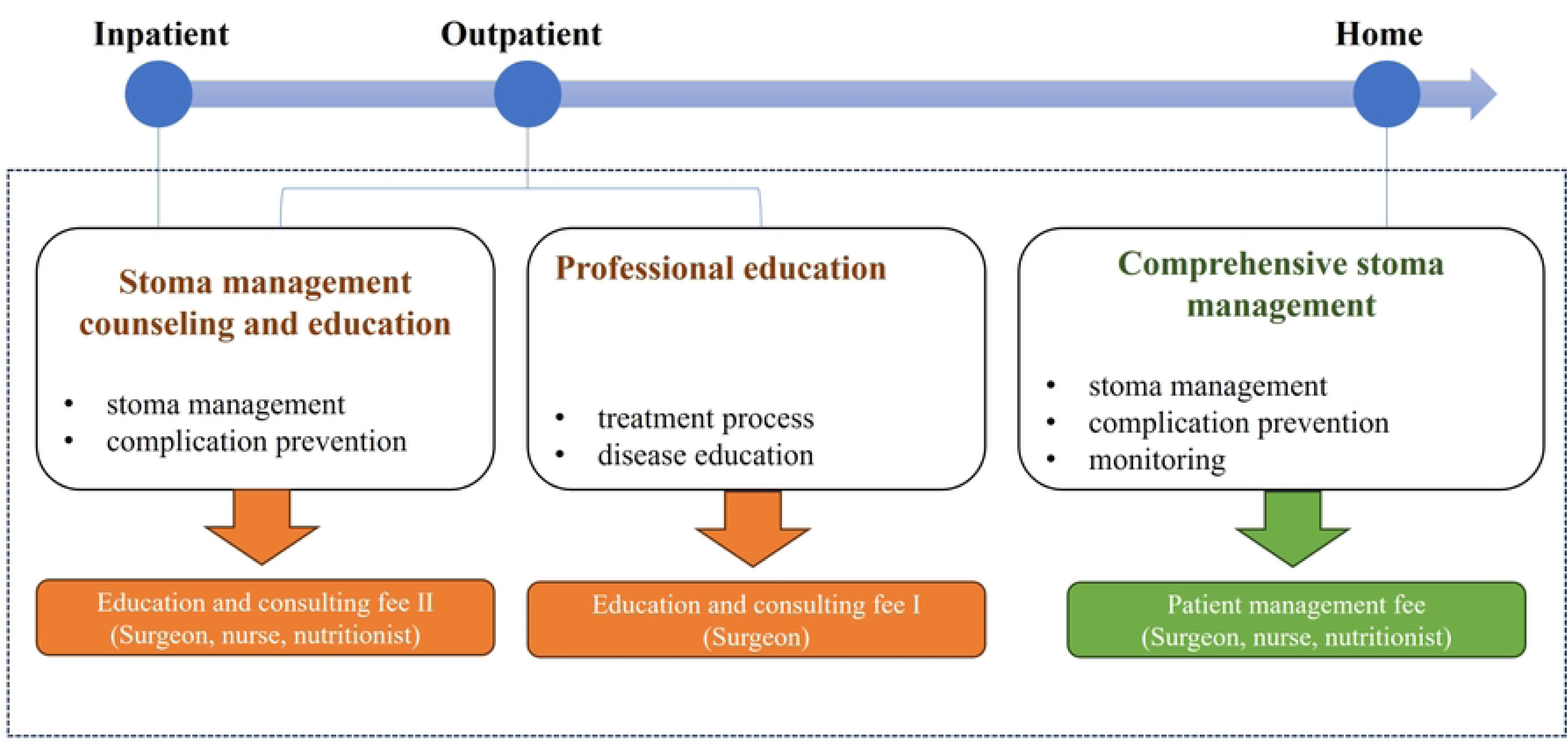
**Overview of the Cancer Patient Home-Care Pilot Program.**

### Study design and sample

A descriptive survey was conducted of Korean Association of Wound Ostomy Continence Nurses to assess their satisfaction and needs regarding participation in the National Cancer Patient Home-Care Pilot Program. The survey covered the period from November 13, 2022, to January 19, 2023, and included nurses with experience in stoma care regardless of their certification as wound, ostomy, or continence nurses and their participation in the pilot project. All participating nurses signed an informed consent form before the survey, and the collected data were processed anonymously.

To estimate the minimum sample size for a 95% confidence level and a 5% sample error, the number of wound, ostomy, and continence nurses in South Korea was estimated to be approximately 3,000. However, because of restrictions related to nurses currently not practicing, the number of nurses participating in the pilot project, data collection period, and budget, only 207 participants were enrolled in the present study. After excluding 11 participants with incomplete responses, 196 participants were included in the final analysis.

### Measurements

*The following information was elicited through the questionnaire:*

1. General characteristics of respondents, encompassing respondents’ status as a wound, ostomy, or continence nurse and their participation in the home-care pilot project.
2. Current status of management of patients with an ostomy, encompassing the types of educational materials currently used and those that will be needed in the future, as well as the types of contents of these educational materials, for the management of patients with an ostomy.
3. Home Care Pilot Project-related factors, encompassing the number of personnel involved in the home care pilot project, the current status of patients, satisfaction with the pilot project fee system, satisfaction with the pilot project, needs of the nurses, and changes in patient care and consultations.

### Ethical considerations

This study was approved by the Institutional Review Board of the NCC Center (IRB No.: NCC2022-0849). The informed consent form contained information about the study background, purpose, procedures, method, sample size, participation period, anticipated benefits and risks/inconveniences for study participation, withdrawal of consent, and disclosure and protection of personal information. Additionally, the researchers’ contact information was provided so that the participants could contact the researchers at any time regarding any study-related questions. Both the survey and informed consent forms were collected in written form.

### Data analysis

The data collected in the study were analyzed using R software (version 4.2.1; University of Auckland, Auckland, New Zealand). Study participant characteristics and survey results were analyzed using descriptive statistics, and the differences in the types of educational materials currently used and needed for stoma care education according to participation in the pilot project were analyzed using the chi-square test. A p-value >0.05 was considered statistically significant.

## Results

### Current status of the National Cancer Patient Home-Care Pilot Program

Of the 196 participants, 42 (21.4%) participated in the home-care pilot program for cancer patients with an ostomy, and 29 (69.0%) were certified wounds, ostomy, and continence nurses. Sixteen (38.1%) nurses participating in the pilot project joined in 2021, while 21 (50.0%) had joined in 2022. The average number of physicians, nurses, and nutritionists in the home-care team was 3.8, 3.1, and 4.8, respectively. The nurses who participated in the survey provided home-care services to an average of 158.7 patients. Of the 42 nurses currently participating in the home-care pilot project, 35 (85.4%) were satisfied with the project overall, while 6 (14.6%) were neither satisfied nor dissatisfied. Thirty-eight (92.7%) of them were willing to continue participating in the National Cancer Patient Home-Care Pilot Program, and three (7.3%) were undecided. Table 1 reported the types of materials used by nurses who participated in the pilot project and nurses who did not participate in educating patients with stoma and the types of materials required to educate patients with stoma. Regarding the educational materials used, hospital-developed printouts were the most common type used to educate cancer patients with an ostomy (90.5%), followed by videos (76.2%), stoma models (60.0%), and printouts provided by a stoma-related company (27.0%). The most common educational materials used by nurses not participating in the pilot project were hospital-developed printouts (50.0%), followed by hospital-developed videos (42.0%), printouts provided by a stoma-related company (49.1%), and stoma models (40.3%). Hospital-developed printouts and videos were more commonly used by nurses currently participating in the pilot project (p<.001 and p<.001), whereas videos provided by a stoma-related company were less commonly used (p=.025).

**Table 1.** Types of Educational Materials Used for Education of Cancer Patients with Stoma and Types of Educational Materials Needed.

| Variables | Nurse who participated in a pilot project, n (%) |  |  | Nurse who does not participate in a pilot project, n (%) |  |  | *P* |
| --- | --- | --- | --- | --- | --- | --- | --- |
|  | Mainly used | Used as an aid or when necessary | Do not use | Mainly used | Used as an aid or when necessary | Do not use |  |
| **Educational materials used** |  |  |  |  |  |  |  |
| Printed material developed by the hospital | 38 (90.5) | 2 (4.8) | 2 (4.8) | 77 (55.0) | 48 (34.3) | 15 (10.7) | <.001 |
| Printed material developed by the society | 10 (27.0) | 20 (54.1) | 7 (18.9) | 35 (26.5) | 73 (55.3) | 24 (18.2) | .990 |
| Printed material provided by the company related to the stoma | 10 (27.0) | 17 (45.9) | 10 (27.0) | 56 (40.3) | 64 (46.0) | 19 (13.7) | .104 |
| Video materials developed by the hospital | 32 (76.2) | 1 (2.4) | 9 (21.4) | 58 (42.0) | 52 (37.7) | 28 (20.3) | <.001 |
| Video materials developed by the society | 7 (19.4) | 16 (44.4) | 13 (36.1) | 25 (18.9) | 70 (53.0) | 37 (28.0) | .597 |
| Video material provided by the company related to the stoma | 7 (19.4) | 14 (38.9) | 15 (41.7) | 42 (30.0) | 70 (50.0) | 28 (20.0) | .025 |
| Stoma model | 21 (60.0) | 10 (28.6) | 4 (11.4) | 56 (49.1) | 40 (35.1) | 18 (15.8) | .523 |
| **Educational materials needed** | Necessary | Average | Not necessary | Necessary | Average | Not necessary | *P* |
| Printed material developed by the hospital | 41 (97.6) | 1 (2.4) | 0 (0.0) | 127 (88.2) | 13 (9.0) | 4 (2.8) | .183 |
| Printed material developed by the society | 36 (85.7) | 5 (11.9) | 1 (2.4) | 124 (84.4) | 18 (12.2) | 5 (3.4) | .943 |
| **Printed material provided by the company related to the stoma** | **21 (52.5)** | **11 (27.5)** | **8 (20.0)** | **114 (78.1)** | **28 (19.2)** | **4 (2.7)** | **<.001** |
| **Video materials developed by the hospital** | **37 (88.1)** | **5 (11.9)** | **0 (0.0)** | **119 (81.0)** | **23 (15.6)** | **5 (3.4)** | **.379** |
| **Video materials developed by the society** | **35 (83.3)** | **6 (14.3)** | **1 (2.4)** | **122 (83.6)** | **17 (11.6)** | **7 (4.8)** | **.729** |
| **Video material provided by the company related to the stoma** | **21 (52.5)** | **15 (37.5)** | **4 (10.0)** | **118 (80.8)** | **22 (15.1)** | **6 (4.1)** | **.001** |
| **Stoma model** | **36 (90.0)** | **3 (7.5)** | **1 (2.5)** | **119 (84.4)** | **19 (13.5)** | **3 (2.1)** | **.592** |
| **XR-based digital contents** | **30 (78.9)** | **7 (18.4)** | **1 (2.6)** | **100 (73.5)** | **27 (19.9)** | **9 (6.6)** | **.616** |

Regarding the types of educational materials needed to educate cancer patients with an ostomy, the nurses participating in the pilot project reported that hospital-developed printouts (97.6%), stoma models (90.0%), hospital-developed videos (88.1%), printouts developed by stoma-related academic societies (85.7%), videos developed by stoma-related academic societies (83.3%), and extended reality (XR)-based digital content (78.9%) were the most needed. The nurses who did not participate in the pilot project reported that hospital printouts (88.2%), printouts developed by stoma-related academic societies (84.4%), stoma models (84.4%), videos developed by stoma-related academic societies (83.6%), videos developed by the hospital (81.0%), and videos provided by a stoma-related company (80.8%) were the most needed. The need for printouts and videos provided by a stoma-related company was significantly higher among nurses who did not participate in the pilot project.

### Evaluation of the effectiveness of the National Cancer Patient Home-Care Pilot Program

Table 2 reported on the evaluation of the effectiveness national cancer patient home-care pilot program. Thirty-nine (92.9%) nurses reported that the pilot program helped improve the quality of patient care, while 38 (90.5%) nurses mentioned that the pilot program helped cancer patients to understand ostomies. Thirty-nine (92.9%0 nurses stated that the program helped build a trusting relationship between healthcare professionals and patients, while 29 (69.0%) mentioned that the number of inquiries about simple health-related information after a hospital appointment decreased. Regarding changes in the healthcare professionals who participated in the pilot project, 40 (95.2%) mentioned that they were able to provide easier and more detailed explanations to the patients, and 40 (95.2%) mentioned that they listened to the patients more and reflected their opinions in the treatment process. Thirty-nine (92.9%) stated that they now assessed patients’ clinical problems more comprehensively and reflected them in treatment, and 39 (92.9%) stated that they were able to build a more trusting relationship with the patients. Forty (95.2%) stated that patient satisfaction with patient care increased, while 41 (97.6%) reported that the program helped enhance their expertise.

**Table 2.**
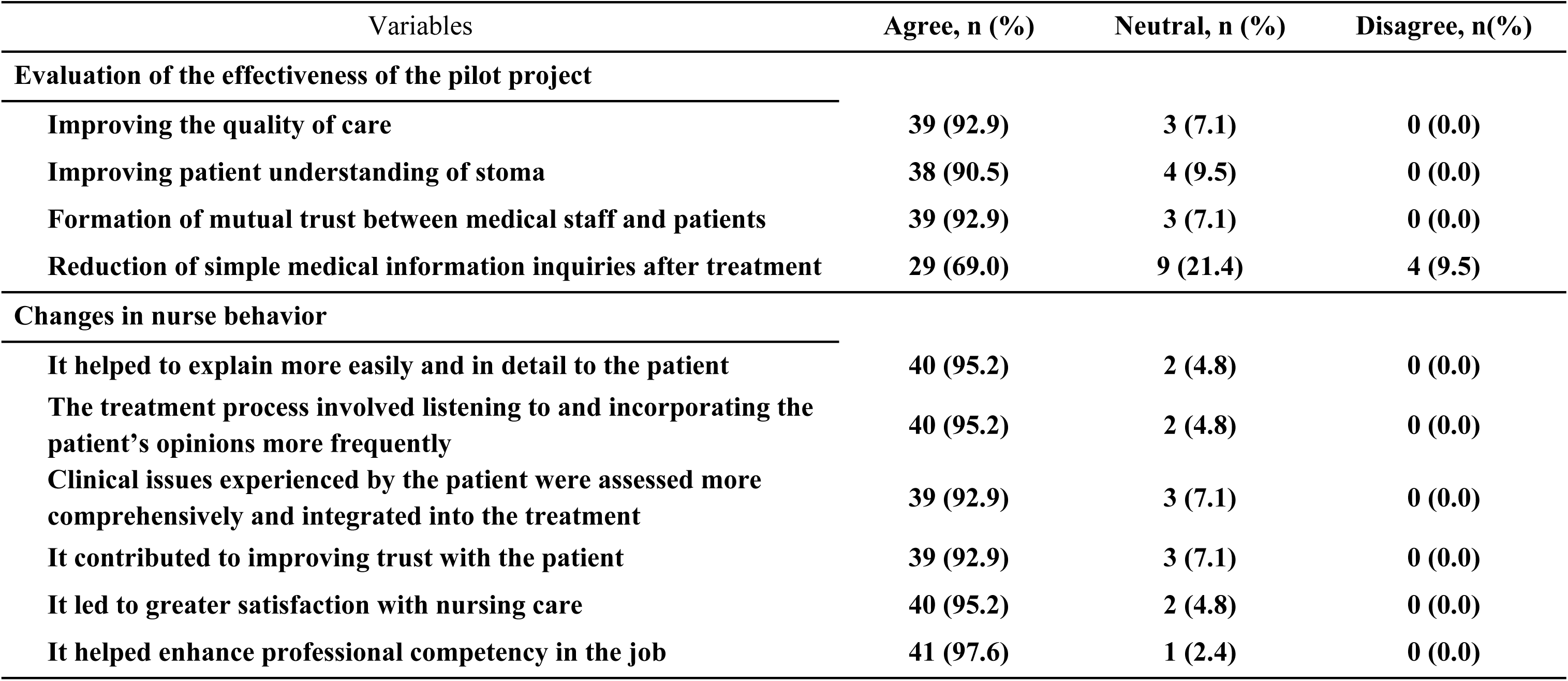
Evaluation of the Effectiveness of the Pilot Project for At-Home Medical Care for Cancer Patients with Stoma and Changes in Nurse Behavior.

### Evaluation of the fee system for the National Cancer Patient Home-Care Pilot Program

Table 3 reported on the evaluation of the fee system for the national cancer patient home-care pilot program. Satisfaction with the fee system of the National Cancer Patient Home-Care Pilot Program among nurses currently participating in the program was assessed. Nineteen (45.2%) nurses did not agree to the “Education and Consultation fee II: limited to 6 times a year,” but 25 (61.0%), 23 (54.8%), and 27 (64.3%) agreed to the criteria for “Education and Consultation fee II: ≥20 minutes each session,” “Education and Consultation fee II: fee system,” and “Education and Consultation fee II: out-of-pocket cost,” respectively. Twenty-five (61.0%) nurses did not agree to the criterion “Patient management fee: reimburse 1 session when 2 sessions are provided a month,” and 19 (48.7%) agreed to the criterion “Patient management fee -fee system”.

**Table 3.** Satisfaction with Cancer Patients’ Home Medical Treatment Pilot Project Medical Fee Criteria.

| Variables | Agree, n (%) | Neutral, n (%) | Disagree, n (%) |
| --- | --- | --- | --- |
| **Education and consulting fee II** |  |  |  |
| Limited to 6 times a year | 15 (35.7) | 8 (19.0) | 19 (45.2) |
| At least 20 minutes each time | 25 (61.0) | 11 (26.8) | 5 (12.2) |
| Medical fee | 23 (54.8) | 10 (23.8) | 9 (21.4) |
| Co-insurance | 27 (64.3) | 11 (26.2) | 4 (9.5) |
| **Patient management fee** |  |  |  |
| Recognized once every 2 times a month | 10 (24.4) | 6 (14.6) | 25 (61.0) |
| Medical fee | 19 (48.7) | 11 (28.2) | 9 (23.1) |

### Measures to improve National Cancer Patient Home-Care Pilot Program

Thirty (71.4%) nurses reported that the number of education and consultation sessions provided and the duration of these sessions should be improved. In addition, 27 (64.3%) mentioned that additional personnel, such as nurses and social workers, are needed in home-care teams, while 23 (54.8%) stated that the administrative process and inspection forms must be streamlined. Twenty-five (59.5%) nurses who were currently participating in the pilot project stated that they could not provide education and consultation or patient management for cancer patients, and the major reasons were shortage of healthcare service providers and patients’ hospitalization in another healthcare facility.

Among nurses not currently participating in the National Cancer Patient Home-Care Pilot Program, the major reasons for not participating in the program were shortage of personnel or lack of facilities (n=34, 23.5%), and lack of promotion of the program (n=34, 23.5%).

## Discussion

Cancer patients who have an ostomy must make adjustments to stoma care and physical changes related to the stoma. These adjustments can have a significant impact on their physical, psychological, and social well-being. Failure to properly care for a stoma can result in complications, anxiety, depression, and a loss of self-esteem due to physical changes, which can lead to physical and psychological health problems [15]. Patients who receive stoma care education are able to adjust to their physical changes more quickly and easily and care for their stoma more skillfully, along with alleviated depression and anxiety [16]. Therefore, this study investigated the current status and effectiveness of the National Cancer Patient Home-Care Pilot Program and measures for improvement through a survey of nurses currently participating in the program and nurses not participating in the program but who had experience with stoma care education and practice. The ultimate goal was to promote the National Cancer Patient Home-Care Pilot Program and facilitate the transition of the pilot program into a fully operational program so that adequate healthcare services are provided to cancer patients with an ostomy.

The nurses showed a high level of satisfaction with the National Cancer Patient Home-Care Pilot Program overall, and none of them claimed to be dissatisfied with the program. In addition, the nurses mentioned that the pilot program helped improve the quality of patient care, helped patients understand their ostomies, and helped build a trusting relationship between healthcare professionals and patients. The results of this study support previous findings of a systematic review that continuous monitoring after discharge and continuous management by an ostomy nurse is significantly more effective in improving health outcomes and treatment satisfaction than providing stoma care education only before discharge [17]. In other words, patients with an ostomy can properly manage their stoma and peristomal skin and resolve their physical, social, and psychological problems caused by the stoma through continuous monitoring after discharge [18].

The services that can be provided to cancer patients with an ostomy by nurses as a part of the National Cancer Patient Home-Care Pilot Program are “Education and Consultation II” and “Patient Management.” “Education and Consultation II” services include education and consultation provided by the home-care team for patients visiting the hospital and their caregivers regarding stoma care, prevention and management of stoma complications, diet, management of activities of daily living, and mental health management following discharge. “Patient Management” services include periodic monitoring of stoma functioning and health by the home-care team and confirming patients’ understanding of the provided education and providing repeated education through a virtual bidirectional service as a part of the “Education and Consultation” service. The reimbursement criteria for “Education and Consultation II” services include individual education provided to patients and caregivers for at least 20 minutes a session, with up to one session per day and six sessions per year reimbursed. The reimbursement criteria for “Patient Management” services include one session per month (from the first day to the last day of the month) during a one-year period only when at least two virtual patient management (bidirectional) services are provided in the same month [14]. However, the greatest need among nurses participating in the National Cancer Patient Home-Care Pilot Program pertained to the fee system; 71.4% of the nurses participating in the program mentioned that the current fee system must be improved. Although up to six education and consultation sessions provided by the home-care team to a single patient can be reimbursed, the nurses stated that additional education and consultation sessions may be needed for patients with other comorbidities, those who developed complications, and those who required more time to adjust to the ostomy. In addition, the nurses mentioned that the reimbursement criteria for patient management services (reimbursement offered for one session per month only when a minimum of two sessions are performed in a given month) are highly restrictive in actual clinical settings. For instance, the current criterion defines a month as the period from the first day of a calendar month to the last day of the month, so if the first service is provided near the end of the month, another session must be performed as soon as possible to receive reimbursement for one of the sessions. Moreover, some patients are discharged to a long-term care hospital; therefore, even when one session of patient management service is provided to the patient, additional sessions cannot be provided because of their admission to a long-term care hospital, which then disqualifies the first session for reimbursement. Therefore, the current criterion limited to reimbursement of up to six sessions per year should be revised in consideration of factors such as patient complications, comorbidities, type of ostomies, and level of comprehension of stoma care education. In addition, the current criteria for the reimbursement of patient management services, wherein reimbursement is offered only when at least two sessions of services are provided in a month, should be revised to allow reimbursement even when only one session is provided in a month.

The primary reasons for the current lack of participation in the National Cancer Patient Home-Care Pilot Program were a shortage of available personnel and a lack of facilities. While an advanced practice nurse system was developed to address the shortage of healthcare providers, it has only partially resolved this issue. However, these nurses are performing their duties without adequate education, training, or support from legislation and systems [19]. Moreover, education systems to cultivate advanced practice nurses are insufficient, which has resulted in a limited availability of such nurses [20]. Experienced and expert nurses were essential to implement the National Cancer Patient Home-Care Pilot Program. Consequently, maintaining nurses’ expertise through stoma care training and fostering more advanced practice would enhance the performance of the National Cancer Patient Home-Care Pilot Program.

A limitation of this study is that we did not survey the satisfaction and needs of the patients and physicians who participated in the National Cancer Patient Home-Care Pilot Program, which is a direction for future studies. Additionally, as this study was based on a questionnaire survey, we should evaluate the effectiveness of the pilot program through other means to improve the pilot project and facilitate its transition to a fully operational program.

## Conclusion

To assess the current status and identify potential improvements for the National Cancer Patient Home-Care Pilot Program, we conducted a questionnaire survey among nurses experienced in stoma care. The results showed high satisfaction with the program and its effectiveness. The nurses also suggested enhancing the reimbursement system and fostering more qualified personnel to expand the program’s application. Therefore, the number of sessions and reimbursed sessions of home-care services should be adjusted, and qualified personnel should be fostered through quality education and training to promote the National Cancer Patient Home-Care Pilot Program and facilitate its transition to a fully operational one.

## Data Availability

Data are available from the National Cancer Center Ethics Committee for researchers who meet the criteria for access to confidential data.

## Acknowledgements

The authors would thank to all the nurses who participated in the survey, and to the Korean association of wound ostomy continence for their help in the survey.

